# Uncertainty quantification in epigenetic clocks via conformalized quantile regression

**DOI:** 10.1101/2024.09.06.24313192

**Authors:** Yanping Li, Jaclyn M. Goodrich, Karen E Peterson, Peter X-K Song, Lan Luo

## Abstract

DNA methylation (DNAm) is a chemical modification of DNA that can be influenced by various factors, including age, the environment, and lifestyle. An epigenetic clock is a predictive tool that measures biological age based on DNAm levels. It can provide insights into an individual’s biological age, which may differ from their chronological age. This difference, known as the epigenetic age acceleration, may reflect health status and the risk for age-related diseases. Moreover, epigenetic clocks are used in studies of aging to assess the effectiveness of anti-aging interventions and to understand the underlying mechanisms of aging and disease. Various epigenetic clocks have been developed using samples from different populations, tissues, and cell types, typically by training high-dimensional linear regression models with an elastic net penalty. While these models can predict mean biological age based on DNAm with high precision, there is a lack of uncertainty quantification which is important for interpreting the precision of age estimations and for clinical decision-making. To understand the distribution of a biological age clock beyond its mean, we propose a general pipeline for training epigenetic clocks, based on an integration of high-dimensional quantile regression and conformal prediction, to effectively reveal population heterogeneity and construct prediction intervals. Our approach produces adaptive prediction intervals not only achieving nominal coverage but also accounting for the inherent variability across individuals. By using the data collected from 728 blood samples in 11 DNAm datasets from children, we find that our quantile regression-based prediction intervals are narrower than those derived from conventional mean regression-based epigenetic clocks. This observation demonstrates an improved statistical efficiency over the existing pipeline for training epigenetic clocks. In addition, the resulting intervals have a synchronized varying pattern to age acceleration, effectively revealing cellular evolutionary heterogeneity in age patterns in different developmental stages during individual childhoods and adolescent cohort. Our findings suggest that conformalized high-dimensional quantile regression can produce valid prediction intervals and uncover underlying population heterogeneity. Although our methodology focuses on the distribution of measures of biological aging in children, it is applicable to a broader range of age groups to improve understanding of epigenetic age beyond the mean. This inference-based toolbox could provide valuable insights for future applications of epigenetic interventions for age-related diseases.

## 1 BACKGROUND

While chronological age is arguably a strong risk factor for aging-related death and diseases, individuals of the same chronological age may exhibit great heterogeneity in physiologic functions and rate of biological aging. Identifying aging biomarkers is a crucial step in the evaluation of interventions aimed at promoting healthier aging. Epigenetic age is a biomarker of aging that has been reported to be associated with age-related disease and all-cause mortality ^1,2,3,4^. It has been found that composite measures of DNA methylation (DNAm) levels across specific sets of cytosine-phosphate-guanine (CpG) sites, often called epigenetic clocks, are strongly associated with chronological age or age-related diseases. One of the first and most widely used epigenetic age predictors is Horvath’s epigenetic clock ^1^, a statistical prediction model that uses DNAm at 353 CpG sites to predict chronological age.

Standard approaches for training epigenetic clocks involve several key steps: (i) collecting biological samples from individuals with diverse backgrounds; (ii) extracting DNA and performing DNA methylation analysis; (iii) conducting data preprocessing procedures such as missing data imputation, outliers removal, and data normalization; (iv) adopting a feature screening method for identifying relevant CpG sites that are predictive of age or linked with aging processes; (v) fitting a high-dimensional regression model with elastic net penalty; and (vi) evaluating model performance on an independent test dataset to verify its accuracy and robustness.

Despite the well-established pipelines for constructing epigenetic clocks, most of them only provide point mean predictions ^1,2,5^. Recently, there has been work focusing on the construction of confidence intervals for DNAm age using a novel U-learning approach ^6^. While their method is valuable for quantifying uncertainty in parameter estimation, it differs from our goal of constructing prediction intervals that include both variability of sample points and parameter uncertainty. In biological age prediction, it is important not only to predict accurately but also to quantify the uncertainty of the predictions. This is especially true in biological systems that are inherently complex and exhibit variability both within and between individuals. Uncertainty quantification helps to assess the reliability of the models used to construct these clocks, accounting for the variability and stochastic nature of biological aging processes. The uncertainty in a prediction can be quantified using a prediction interval, giving lower and upper bounds between which the response variable lies with high probability. More specifically, epigenetic clocks aim to measure biological age, which can differ significantly from chronological age due to various factors such as lifestyle, genetics, and the environment. Prediction intervals help to interpret these predictions by indicating how much variation there is around the estimated biological age, thus helping to better understand the potential impacts of these factors. In addition, when clinicians or researchers use epigenetic clocks to study aging or assess the risk of age-related diseases, prediction intervals can help them assess the reliability of the clock in different populations or under different conditions. A narrow interval suggests high reliability and vice versa, influencing the confidence in using these tools for medical or research decisions. However, it is worth noting that existing epigenetic clocks are primarily built from ultra high-dimensional DNAm data where most inferential methods rely on nontrivial assumptions such as the linear model being true, the error distribution, the homoscedasticity of errors, etc. ^7,8,9,10^.

In high-dimensional settings, where the number of predictors (CpGs) exceeds the number of subjects, traditional statistical methods often struggle to provide reliable uncertainty estimates due to overfitting and high variance. Conformal prediction methods can mitigate these issues by constructing prediction intervals that are valid under a minimal set of assumptions about the data distribution ^11^. They can be applied on top of any predictive model, and this flexibility is particularly useful in constructing prediction intervals for epigenetic clocks. A key component of conformal prediction is the nonconformity score, which quantifies how well a new data point conforms to previously observed data. In the study of epigenetic aging, one of the natural choices of the nonconformity score can be the residual from regressing an epigenetic clock on chronological age, referred to as epigenetic age acceleration which occurs when an individual’s DNAm age exceeds their chronological age. Prior studies suggest that older epigenetic age may be associated with lower levels of physical functioning, and declines in global cognitive functioning among long-lived individuals ^3,4,12,13,14^. Since epigenetic age acceleration also has been found to be related to adverse outcomes such as cardiovascular diseases and cancer, it is also scientifically meaningful to incorporate this important biological measure in constructing age prediction intervals. However, commonly used high-dimensional linear regression models for building existing epigenetic clocks implicitly assume that the association between the DNAm profile and chronological age remains the same across different subpopulations, which may not hold due to heterogeneity across different developmental stages, health conditions, and genders ^5^. Moreover, constructing prediction intervals from conditional mean regression results in intervals of uniform width, failing to capture individual variability in aging rates or age acceleration, which are influenced by many factors including sociodemographic factors, diet, physical activity, genetics, environmental chemical exposures, and more. Apparently, prediction intervals of the same width, even if statistically valid with nominal coverage, are not able to capture cross-individual heterogeneity in the age acceleration rate across different risk levels.

To bridge this gap, we are interested in looking into subgroup differences across various factors that may affect age acceleration and incorporating such variability into age prediction intervals. First, we propose to build a quantile regression framework for constructing epigenetic clocks. This would allow researchers to examine relationships between DNAm levels and different quantiles of the chronological age. Apparently, different age groups may show a different pattern in their methylation levels across CpG sites, calling for age-specific epigenetic clocks. For example, we expect that the same CpG site to have varying effects across different age groups. Characterizing such information can provide insight on the impact of DNAm on different age groups or health outcomes. Furthermore, we plan to construct age prediction intervals within the quantile regression framework.

For example, to obtain prediction intervals with 90% nominal coverage, we simply fit the conditional quantile function at the 5% and 95% levels to form the corresponding intervals. This method is robust to data with high heteroscedasticity and adaptive to local variability. Besides providing valid coverage in finite samples, the intervals are as short as possible with their length adaptive to individual variability. This enables the identification of individual characteristics that are associated with different levels of uncertainty in their outcome predictions, facilitating tailored interventions.

Throughout this article, we will use children’s DNAm data from the Gene Expression Omnibus (GEO) database to illustrate our proposed pipeline for constructing prediction intervals for biological age. Similar to the first-generation epigenetic clocks, we will use chronological age as the outcome of interest. Previous works have adopted a high-dimensional linear regression model with elastic net penalty ^15^, see for example, the Horvath clock ^1^, Hannum clock ^2^, and Levine clock ^3^. However, due to the underlying heterogeneity across different age groups, the associations between DNAm levels and age may vary. As a comparison to the conventionally used high-dimensional linear regression model, we will first fit a high-dimensional quantile regression model with the elastic net penalty. Then we will construct prediction intervals and demonstrate their validity as well as statistical efficiency. Our results show that the median regression model outperforms the mean regression model, exhibiting a higher correlation between chronological and predicted ages and a lower median error. Furthermore, our newly proposed approach for constructing prediction intervals not only accommodates subpopulation heterogeneity but is also statistically more efficient than the conventional mean regression-based method.

We describe the generic pipeline of prediction intervals construction for epigenetic clocks in Section 2. Application of the pipeline to establish epigenetic clocks with the children’s dataset is given in Section 3. Finally, in Section 4, we present a discussion, outline the limitations, and suggest areas for future research.

## 2. METHODS

The overarching goal of this work is to construct prediction intervals for epigenetic clocks with adaptive widths that account for cross-subjects variability. Our proposed generic pipeline involves the following two major components: (i) training high-dimensional predictive models for predicting epigenetic ages, and (ii) constructing valid prediction intervals with the conformal inference methods. A diagram of the proposed generic pipeline is shown in Figure 1.

**FIGURE 1.**
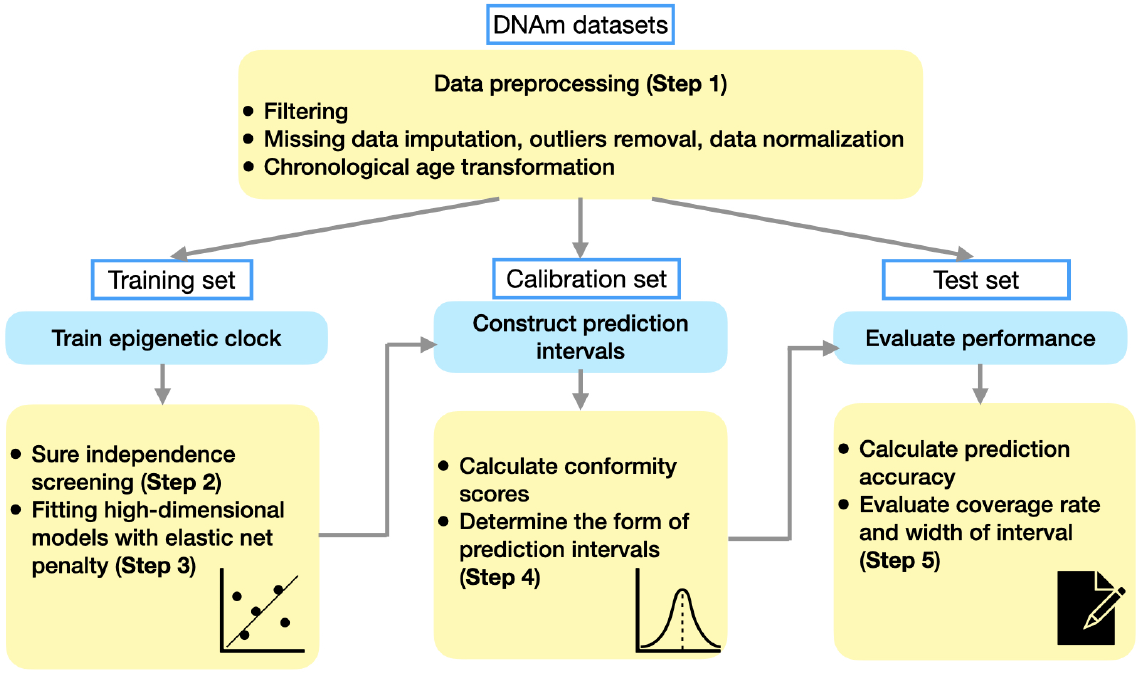
Flowchart of the proposed generic pipeline that integrates two key components: (i) training epigenetic clock, (ii) constructing prediction intervals, and (iii) evaluating performance as highlighted in the blue boxes. Details of each step (shown in yellow boxes) will be presented in Section 2.

### 2.1 Quantile regression based epigenetic clock

DNA methylation data is typically characterized by ultra-high dimensions since hundreds of thousands of CpG sites are profiled. The entire procedure to construct quantile regression based epigenetic clock can be separated into three steps. In our regression problem, we observe independent and identically distributed samples (*X*_*i*_, *Y*_*i*_) ∈ ℝ^*q*^ × ℝ ∼ *P*, where *X*_*i*_ ∈ ℝ^*q*^ is the vector of methylation levels of profiled CpG sites, and *Y*_*i*_ ∈ ℝ is the chronological age of subject *i* for *i* = 1, …, *n*.

#### Step 1 (Data preprocessing)

As mentioned by ^1^and ^5^, we adopt similar data preprocessing methods. In particular, we first integrate data by concentrating on common CpG sites, with several public Illumina DNA datasets relevant to target population. Then we discard any CpG site with more than 10 missing DNA methylation values. For sites with fewer than 10 missing values, we impute with the *k*-nearest-neighbors approach with the `impute` package in `R`. We study overlapping CpGs that are present on all datasets and carry out the normalization step to ensure that these data are comparable by adapting the `BMIQ R` function from ^16^ so that it would rescale all probes of each array to match their distribution with the determined gold standard. We also perform a principal component analysis to identify and remove outliers by converting each sample into a Z-score statistic, transforming it to the false-discovery rate, and removing samples falling below a false-discovery rate of 0.2. To avoid negative values in the predicted age, we apply a transformation function *G*(*Y*) to the original chronological age before model fitting, with the transformed age denoted by 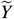. The transformation function *G*(*Y*) should be strictly increasing to preserve the order of ages, invertible to allow mapping back to the original scale, and continuous to ensure smoothness. Additionally, the inverse-transformed values *G*^−1^(*Y*) must be strictly positive to ensure valid age predictions. For example, a logarithmic transformation satisfies all these requirements. After constructing prediction intervals, we will convert them back to their original scale using the exponential function to ensure positivity. Lastly, the entire dataset is randomly split into training set, calibration set and test set with a proper proportion ^17^. Let 𝒟_train_, 𝒟_cal_ and 𝒟_test_ denote the set of sample indices in the training set, calibration set and test set respectively.

#### Step 2 (Feature screening)

Similar to the training procedures for most existing epigenetic clocks, we first adopt the sure independence screening (SIS) method to reduce the number of CpG sites. Since we plan to fit high-dimensional quantile regression models in our subsequent analysis, we choose to work with a model-free generic SIS procedure with fewer and less restrictive assumptions ^18^. For example, correlation-based SIS can be summarized as several sketched stages. Firstly, for each feature *X*_*j*_, compute its correlation with the response variable 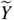, i.e.,

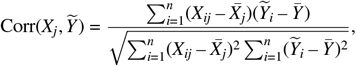

where 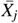 and 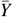 are the mean values of the feature *X*_*j*_ and the response variable 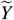, respectively. Then rank the features based on the absolute values of their correlations, i.e.,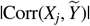. Finally, select the top *p* features with the highest absolute correlations, where *p* is a predetermined number of features to retain. Let 𝒮 ⊂ {1, 2, …, *q*} be the set of the index of selected features, and the size of 𝒮 is obviously *p*. To determine the most appropriate feature screening procedure for the analysis of our dataset, we try several popular screening methods that have been implemented in the `MFSIS` package in `R`, such as SIRS ^19^, DC-SIS ^20^, Kfilter ^21^, CSIS ^18^, Bcor-SIS ^22^, and WLS ^23^ to the training set. For each method, we use a 10-fold cross-validation approach to fit regression models predicting chronological age from the selected CpG sites. Within each fold, we optimize the models by selecting the best tuning parameters and compute the correlation coefficient between predicted values and true values. We evaluate the performances of all different types of feature screening methods across all models in our subsequent analyses, and choose the one that fits best to each of the regression models.

#### Step 3 (High-dimensional quantile regression)

After applying the feature screening method, we then fit a high-dimensional quantile regression model with elastic-net penalty with selected CpGs. It is implemented with the `conquer` package that adopts a convolution-type smoothed quantile regression ^24^. Let 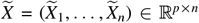 be the matrix that contains only selected features in Step 2. The estimated coefficient vector 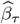 is given by

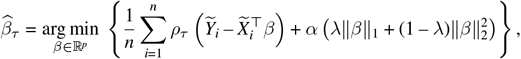

where *ρ*_*τ*_ (*u*) = *u*(*τ* – **1**_{*u*<0}_) is the quantile loss function, ∥*β*∥_1_ and ∥*β*∥_2_ refer to the 𝓁_1_-norm and 𝓁_2_-norm separately of the coefficient vector, *α* > 0 is the overall regularization strength, and 0 ≤ *λ* ≤ 1 is the regularization parameter that controls the balance between 𝓁_1_ (Lasso) and 𝓁_2_ (Ridge) regularization. As a result, the fitted *τ* th conditional quantile of epigenetic age given the methylation levels is formed by 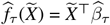, where 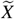 is the vector of DNA methylation level in a new subject.

As a baseline comparison, we also fit a high-dimensional conditional mean regression model with elastic net penalty ^15^ using the `R` package `glmnet`. The corresponding estimated coefficient vector is the solution to the following optimization problem:

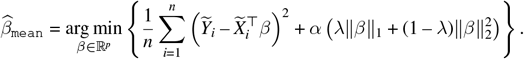

The resulting mean regression based epigenetic clock is given by 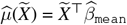.

### 2.2 Uncertainty quantification and interval construction in age prediction

The aforementioned pipeline for constructing epigenetic clocks in Section 2.1 provides point predictions only. However, relying solely on point predictions, without considering the uncertainty or variability, makes it difficult to evaluate the trustworthiness of the predicted biological age or to relate these measures to age-related diseases. Consequently, it is desirable to construct prediction intervals as this quantifies the uncertainty in the prediction and offers a clearer picture of how precise or reliable the age prediction is. While there are various types of conformal prediction methods, we choose to work with the split conformal prediction due to its computational efficiency ^25^. Our goal in this section is to construct a level (1 – *γ*) prediction interval for biological age prediction.

Ideally, we would like to construct prediction intervals that adapt to the heteroscedasticity in the data. This means the resulting intervals can have wider width in subpopulations with high uncertainty and narrow in those of low uncertainty, providing a more accurate depiction of the confidence in age prediction model. Conformalized quantile regression ^17^ merges quantile regression and conformal prediction to produce prediction intervals that adapt to the underlying distribution of the data while maintaining rigorous coverage guarantees. This motivates us to integrate high-dimensional quantile regression models to the conformal prediction framework for epigenetic age predictions.

The framework of split conformal prediction will be integrated with epigenetic clocks based on mean regression and quantile regression, respectively. As a continuation of Section 2.1, we use 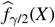 and 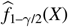 to denote the *γ*/2th and (1 – *γ*/2)th condition quantile functions, respectively. We further use 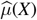 to denote the mean regression based epigenetic clock. The split conformal prediction separates the fitting and ranking steps using sample splitting. More specifically, to avoid overfitting, the fitting or training steps in Section 2.1 is done with the training set 𝒟_train_ while the calibration is carried out with 𝒟_cal_.

#### Step 4 (Residuals and conformity scores)

For the mean regression model, we apply 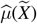 to the samples in calibration set and calculate the absolute values of residuals 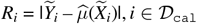. This deviation, in the field of epigenetic aging, is interpreted as the magnitude of age acceleration or deceleration, depending on whether 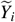 is greater than 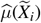. Among the calculated residuals, we find the (1 – *γ*)th quantile of the empirical distribution of the absolute values of residuals, denoted by *d*_mean_ := *d*(ℛ_cal_) = the *k*th smallest value in ℛ_cal_ = {*R*_*i*_ : *i* ∈ 𝒟_cal_}, where *k* = ⌈(*n*/2 + 1) (1 – *γ*)⌉.

Different from the mean regression models, quantile regression offers a natural framework for constructing prediction intervals possibly formed by by 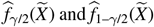. To avoid overfitting, we apply these two fitted quantile functions to the calibration set and calculate the conformity scores defined as 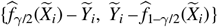, *i* ∈ 𝒟_cal_. Similarly, among the calculated conformity scores, we find the (1 – *γ*)th quantile of the empirical conformity score distribution, denoted by *d*_quantile_ := *d*(𝒮_cal_) the *k*th smallest value in 𝒮_cal_ = {*S*_*i*_ : *i* ∈ 𝒟_cal_}, where *k* = ⌈(*n*/2 + 1) (1 – *γ*)⌉.

#### Step 5 (Constructing prediction intervals)

The prediction interval for the mean regression model is constructed with 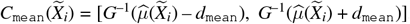. It is clear to see that the conformal prediction technique, if being integrated to the conditional mean regression based epigenetic clock, can be less informative because the resulting prediction intervals are of a relatively consistent width of 2*d*_mean_ across all subjects. More importantly, intervals of approximately constant length do not truly reflect the variation in epigenetic age acceleration across different stages of human lifespan, or more generally, across subpopulations with different health status, gender, ethnicity, and environmental exposures ^26^.

The prediction interval for quantile regression models is constructed with 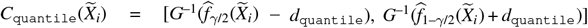, for *i* ∈ 𝒟_test_. Different from the prediction intervals constructed from mean regression based conformal prediction, this method generates intervals with width adaptive to individual DNAm profile and ensures strictly positive lower and upper bounds. In an ideal case where the fitted conditional quantile functions fit perfectly to the calibration set, and *S*_*i*_’s are zero, the prediction interval is simply formed by 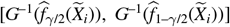 and the prediction intervals will be formed exactly with the two fitted quantile curves. However, if most *S*_*i*_’s are positive and *q*_quantile_ is also positive, the resulting interval will be calibrated to a wider interval, and vice versa. As a result, this way of prediction interval construction automatically adjust for both undercoverage and overcoverage.

### 2.3 Child-specific methylation-based datasets

We acquire several publicly available DNAm datasets from the GEO database, profiled with Illumina 27K and Illumina 450K array platforms (Table 1). We exclude DNAm samples if their chronological ages are missing. These datasets consist of *n* = 728 DNAm samples from healthy children with age ranging from 1 month to 216 months. Such a large cohort of children covers the whole age period from 0 to 18 years helps to study the heterogeneous aging pattern precisely throughout childhood. In addition, utilizing these well-established datasets also ensures a rigorous assessment of its performance, thereby reinforcing the credibility and reliability of our proposed pipeline. These datasets will be instrumental in demonstrating the pipeline designed for developing prediction intervals for epigenetic age, as discussed in Section 3.

**TABLE 1.**
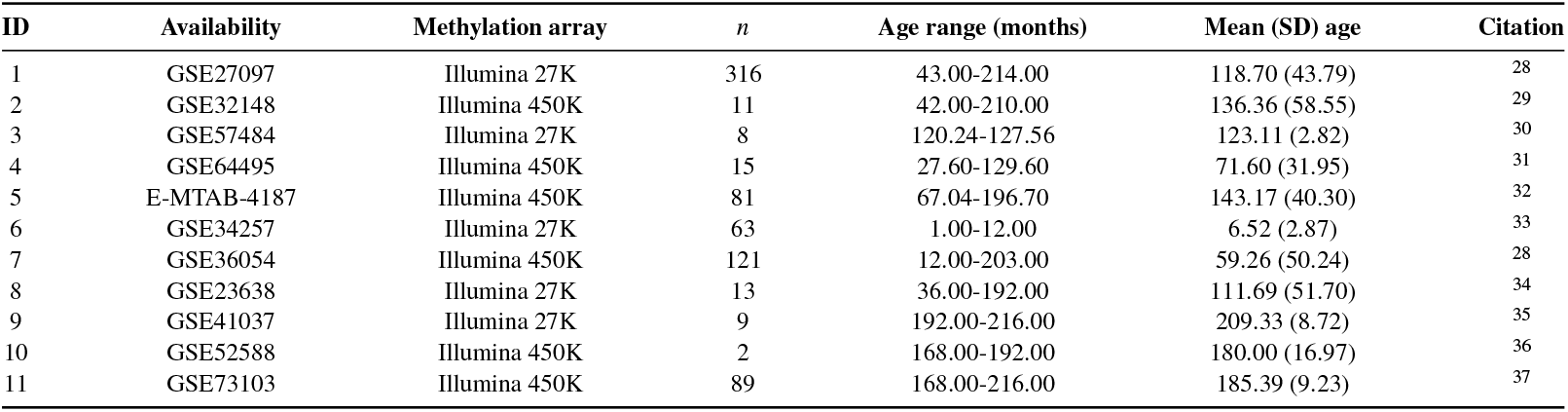
Basic information of the children’s DNA methylation datasets from the GEO database. After preprocessing, the pooled dataset consists of *n* = 728 DNAm samples and *q* = 22, 233 CpG sites.

## 3 RESULTS

In this section, we illustrate the proposed pipeline for constructing prediction intervals for epigenetic age with the children’s datasets. We first introduce the basic characteristics of the DNAm datasets being used. Then we compare the prediction performances between both mean and quantile regression based epigenetic clocks. Lastly, we assess the validity and statistical efficiency of the prediction intervals constructed from these two types of epigenetic clocks, respectively.

### 3.1 Establishment of a child-specific epigenetic clock based on quantile regression

Despite the popularity of conditional mean regression, it is sensitive to outliers and fails to capture heterogeneous relationships between epigenetic age and DNAm profile across different subpopulations. Moreover, in the applications of epigenetic age and aging-related disease predictions, the underlying heterogeneity across subpopulations may not be fully addressed by inferring the conditional mean. As an alternative modeling approach to the conventional linear regression model, quantile regression allows the relationship to vary across quantiles of the chronological age. This helps to uncover the underlying association patterns which may be quite different between children and adults. Basically, quantile regression provides greater flexibility to identify differing relationships at different parts of the distribution of the outcome variable. It has been reported that the variability in epigenetic age differs across human lifespan ^27^. In children and adolescence cohort, for example, the variability in epigenetic age is found to be more drastic in mid-childhood than in toddlers ^5^. For this reason, as an analog of existing epigenetic clocks, we propose to fit a high-dimensional quantile regression model with elastic net penalty.

### 3.2 Characteristics of the DNA methylation datasets (Step 1)

With the children’s datasets, we follow the preprocessing approach illustrated in Step 1 in Section 2.1. Specifically, the gold standard is defined as the mean DNAm values (beta values) from the largest single dataset (GSE27097). This gold standard is used to rescale the probes of all other datasets to ensure comparability, as detailed in Step 1. The age transform function *G*(*Y*) is the same as Function *F* adopted by ^5^, which is defined as:

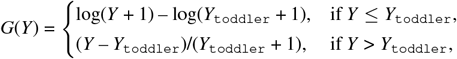

where *Y*_toddler_ is set to 4 years. Then the inverse-transformed function is

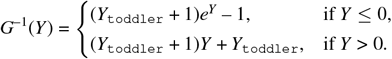

Thus, both *G*(*Y*) and *G*^−1^(*Y*) satisfy the conditions mentioned in Step 1 in Section 2.1: The transformation function *G*(*Y*) is strictly increasing, invertible and continuous, and the inverse-transformed values *G*^−1^(*Y*) is strictly positive. Additionally, the log transformation in the early stages of life, i.e., toddlerhood, reflects the fact that DNAm changes more rapidly during these periods. As individuals grow older, the linear transformation in mid-childhood and adolescence better captures the slower and more gradual changes in DNAm. This approach allows the transformation to adapt to different stages of life in a biologically meaningful way. We apply *G*(*Y*) before building the regression models and use *G*^−1^(*Y*) to transform the predicted values back into the DNAm age. After these steps, our pooled dataset consists of *n* = 728 subjects and *q* = 22, 233 CpG sites. Their ages range from 1 month to 216 months (0 - 18 years). Detailed information is summarized in Table 1.

The entire dataset is randomly split into three sub-datasets: 50% for training set, 30% for calibration set and 20% for test set ^17^. The training set will be used for training a new epigenetic clock while the test set will be used for evaluating the prediction performance of the newly trained clock, see Section 3.3. The hold out calibration set will be used in Section 3.4 for uncertainty quantification and prediction interval construction.

### 3.3 Training a quantile regression based epigenetic clock (Steps 2 to 3)

We first implement a 10-fold cross-validation procedure to determine the number of recruited predictors *p* from the candidate set {[*n*/ log(*n*)], [3*n*/ log(*n*)], [5*n*/ log(*n*)], *n* – 1}, where [*x*] denotes the integer part of *x*. This set is chosen to align with the sure screening property of SIS, as described in Theorem 1 of ^38^. The optimal number of features is selected as *p* = [3*n*/ log(*n*)] = 331. Following Step 2 in Section 2.1 and according to the results summarized in Table 2, we choose the SIRS method for the mean regression model and the Bcor-SIS method for the quantile regression models for dimension reduction because they have the highest overall correlation coefficient within each of the regression models. With these selected CpGs and following Step 3 in Section 2.1, we then fit a high-dimensional median regression model with elastic net penalty. As a baseline comparison, we also fit a high-dimensional mean regression model with elastic net penalty. The mixing parameters *α* are set at 0.15 and 0.45 and the penalty tuning parameters *λ* are set at 0.061 and 0.057 in median (`R` function `conquer.cv.reg`) and mean (`R` function `cv.glmnet`) regression models respectively, based on 10-fold cross-validation of the training data.

**TABLE 2.** After applying different screening methods for dimension reduction in the training sets, we fit various regression models and evaluate their performances by calculating the Pearson correlation coefficients between predicted values and true values. We choose the SIRS method for the mean regression and the Bcor-SIS method for both the 0.05th and 0.95th quantile regression because they have the highest correlation coefficient in each of the regression models.

| Screening methods | Mean regression | 0.05th quantile regression | 0.95th quantile regression | Median regression |
| --- | --- | --- | --- | --- |
| SIRS | **0.92** | 0.88 | **0.90** | 0.90 |
| DC-SIS | 0.91 | **0.89** | 0.89 | 0.90 |
| Kfilter | 0.86 | 0.82 | 0.84 | 0.87 |
| CSIS | 0.91 | 0.88 | 0.89 | 0.90 |
| Bcor-SIS | 0.91 | **0.89** | **0.90** | **0.91** |
| WLS | 0.86 | 0.83 | 0.86 | 0.88 |

To evaluate the prediction accuracy, we apply two different training models to predict ages in the test sets and then compare their Pearson correlation coefficients between DNAm age (predicted age) and chronological age, as well as the median absolute difference between DNAm age and chronological age (median error). As shown in Figure 2, the predictive model based on median regression performs slightly better than the mean regression. We find that the median regression model has a correlation coefficient of 0.886 and a median error of 17 months (Figure 2 (b)), while the correlation coefficient and median error of the mean regression model are 0.878 and 22 months, respectively (Figure 2 (a)). This improvement is largely due to the skewness of the age distribution.

**FIGURE 2.**
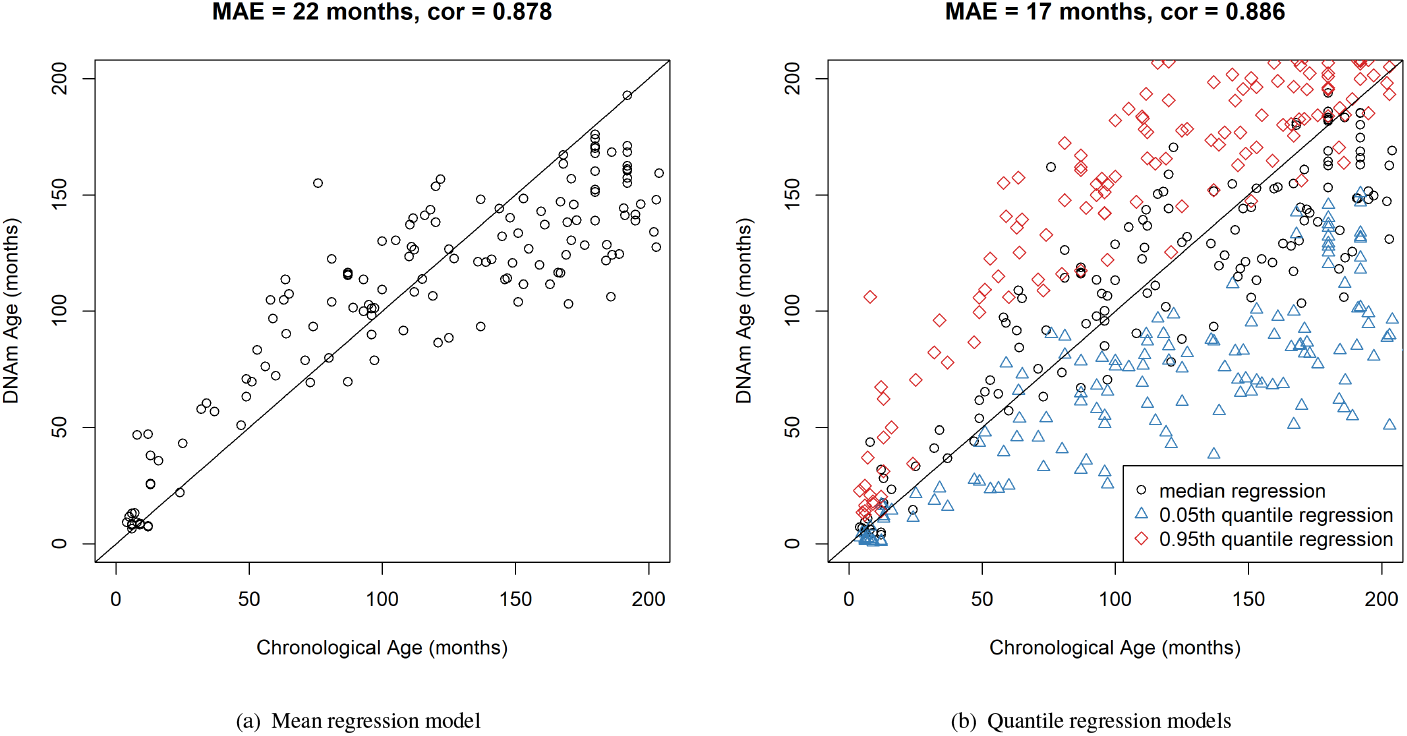
Scatterplots for individuals in the test sets (age unit in month). The straight diagonal lines in both plots are the diagonal references for visualizing the consistency between DNAm age and chronological age. The median regression model (black circles) closely aligns along the diagonal line, indicating a strong correlation with chronological age. Meanwhile, the 0.05th quantile regression model (blue triangles) and the 0.95th quantile regression model (red diamonds) display deviations above and below this line, respectively.

In addition to the median regression model, we also fit the 0.05th and 0.95th high-dimensional quantile regression models to examine the lower and upper tails of the chronological age distributions. In our data example, they correspond to toddlerhood and adolescence, respectively. The mixing parameters *α* are set at 0.31 and 0.23 and the penalty tuning parameters *λ* are set at 0.022 and 0.049 in the 0.05th and 0.95th quantile regression models, respectively, based on a 10-fold cross-validation of the training data. The correspondences between the estimated DNAm and the chronological ages are also plotted in Figure 2 (b). We observe that the blue labels show an upward trend relative to the diagonal reference line. This is because the DNAm ages represented by these blue triangles are the predicted epigenetic ages with a 0.05th quantile regression model, and they represent the estimated threshold for the 5th percentile of the chronological age, conditional on their DNAm levels. Therefore, estimated ages based on DNAm are generally smaller than their true chronological age. Moreover, the deviation from the diagonal line is much smaller for those in the lower tail of the chronological age distribution, while it gradually becomes larger as we move to the upper tail. A similar deviation pattern can be observed from the red diamonds, which show the estimated threshold for the 95th percentile of the chronological age. This phenomenon reveals different variability in epigenetic age across different developmental stages: It varies much less in toddlerhood than in mid-childhood or adolescence. These results demonstrate the benefit and flexibility of fitting high-dimensional quantile regression models as they provide distributional-level knowledge about the epigenetic age.

### 3.4 Constructing prediction intervals and evaluating coverage rate (Steps 4 to 5)

Following Section 2.2, we construct level 90% prediction intervals based on the mean regression and quantile regression models, respectively, by setting *γ* = 0.1. Figure 3(a) shows the prediction intervals derived from the conditional mean regression model. Although they are statistically valid with a coverage rate of 91.1%, which is close to the nominal level, the width of the intervals remains almost the same for all subjects. However, it has been found that there is considerable heterogeneity in physiologic functions and the rate of biological aging among individuals of the same chronological age ^39^. For example, as has been reported in ^5^, age acceleration is the greatest in mid-childhood (5-11 years), and we expect to see wider prediction intervals than those in toddlerhood (0-4 years). Consequently, this way of constructing prediction interval overlooks various heterogeneity across different subpopulations or different developmental stages.

**FIGURE 3.**
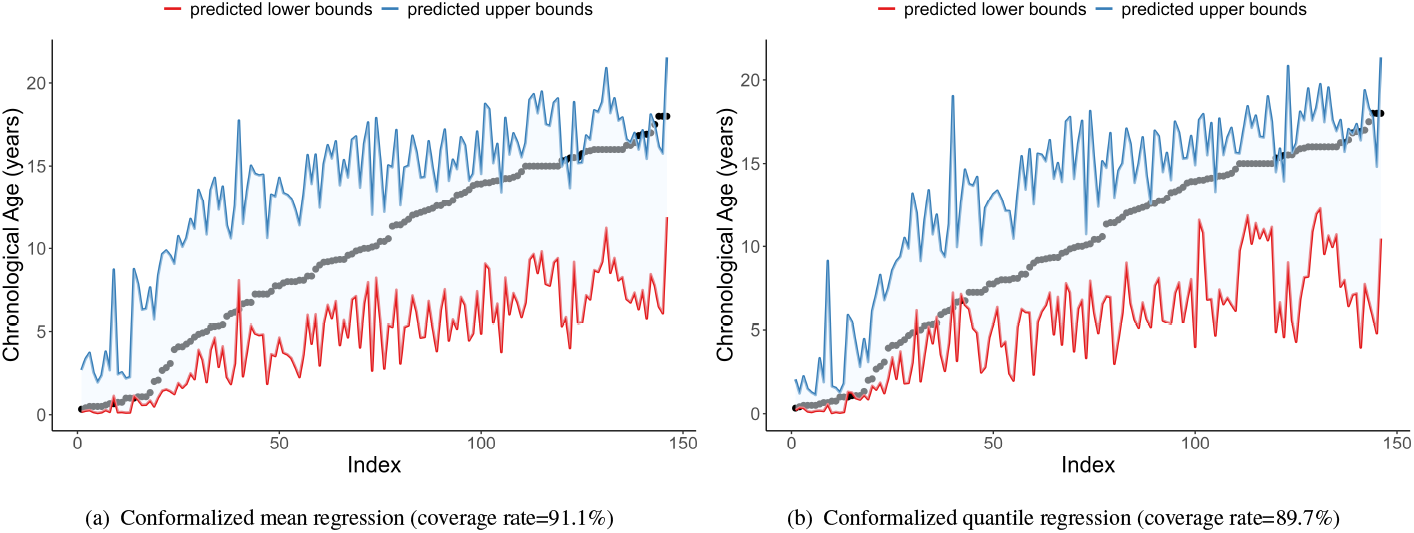
Prediction intervals for individuals in the test sets (age unit in year). The x-axis denotes subjects indexes, and the y-axis represents the true chronological ages. Subject indexes are ordered by chronological ages, as shown in the black dots with an increasing pattern. The blue and red lines denote the upper and lower bounds of prediction intervals, respectively.

In contrast, prediction intervals derived from the quantile regression model are presented in Figure 3(b). With a coverage rate of 89.7% that is also close to the nominal level, the width of the intervals is adapted to individuals with different chronological age. Specifically, we observe that the prediction intervals for toddler age group under 4 years old are much narrower than those of the mid-childhood or adolescence according to Figure 4. Such an observation coincides with that has been reported in ^5^. We further conduct t-test and find that the width of intervals from conformalized quantile method are significantly narrower than those from the mean regression at the significance level *α* = 0.05, with a p-value of 4.357 × 10^−4^. If we stratify by age groups, for subjects in their toddlerhood (p-value = 0.014), mid-childhood (p-value = 1.134 × 10^−5^) and adolescence (p-value = 0.003), the prediction intervals constructed by the conformalized mean regression are all significantly wider than those constructed by the conformalized quantile regression. Besides the width of prediction intervals, comparing the variability of the widths is also of great interest because they reveal the level of heterogeneity across different age groups. We also conduct the Levene’s test to assess whether the variances of the prediction interval widths differ overall and across three age groups. Overall, results show that the variances of the prediction interval widths differ significantly between the two conformal prediction methods at the significance level *α* = 0.05, with a p-value of 0.002. Specifically, in toddlerhood (p-value = 0.451), the variance difference is not statistically significant. However, in mid-childhood (p-value = 7.036 × 10^−16^) and adolescence (p-value = 7.645 × 10^−21^), the variance differences are highly significant. These results suggest that the conformalized quantile regression method captures greater variability in the width of prediction intervals, particularly in the mid-childhood and adolescence stages. Clearly, this approach for constructing prediction intervals provides a more accurate reflection of the heterogeneity in age acceleration across different developmental stages in children.

**FIGURE 4.**
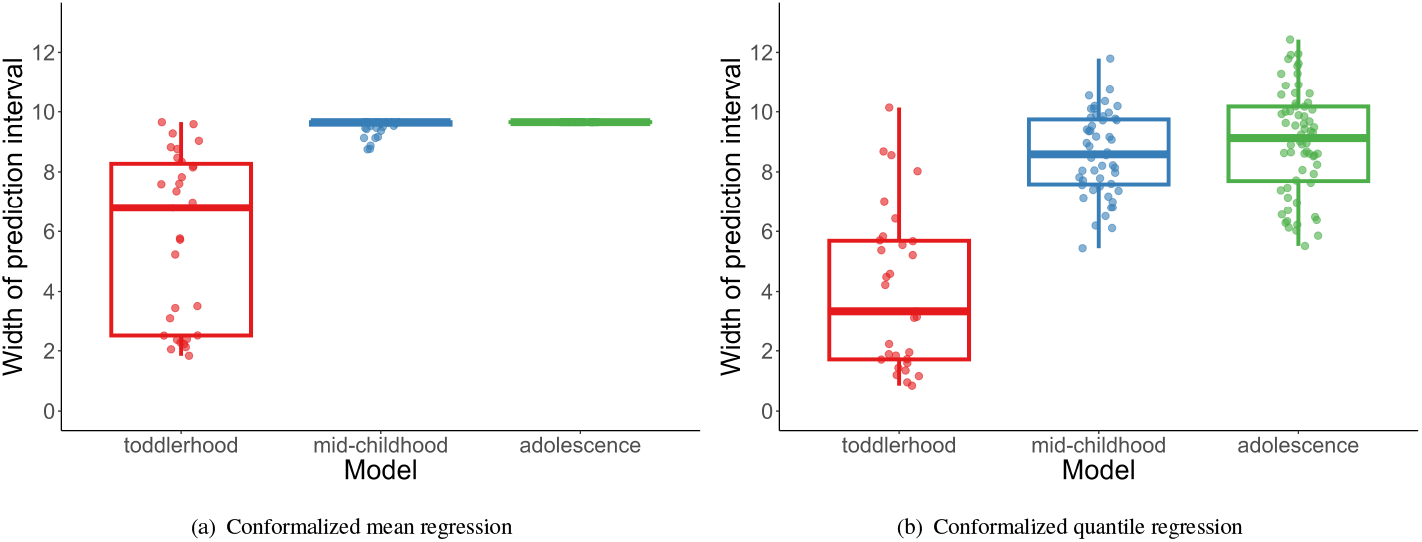
Boxplots of prediction intervals’ width (unit in year) distribution stratified by age groups. The red, blue and green boxplots show the width distributions in toddlerhood, mid-childhood, and adolescence, respectively. The toddler age group has a much narrower prediction intervals than the other two age groups.

With both methods attaining the nominal coverage rate, we further compare their statistical efficiency in terms of prediction interval width. Apparently, efficient statistical methods can result in narrower prediction intervals, reflecting greater certainty and confidence in predictions. As shown in Figure 5, prediction intervals derived from the conformalized quantile regression method are narrower than those approximately constant width intervals produced by conformalized mean regression method: the top of the box on the right, corresponding to the third quartile, is slightly lower than the horizontal line on the left. Consequently, our newly proposed way of constructing epigenetic clocks and their corresponding prediction intervals is not only adaptable to heterogeneity among subpopulations but also statistically more efficient than conventional mean based methods.

**FIGURE 5.**
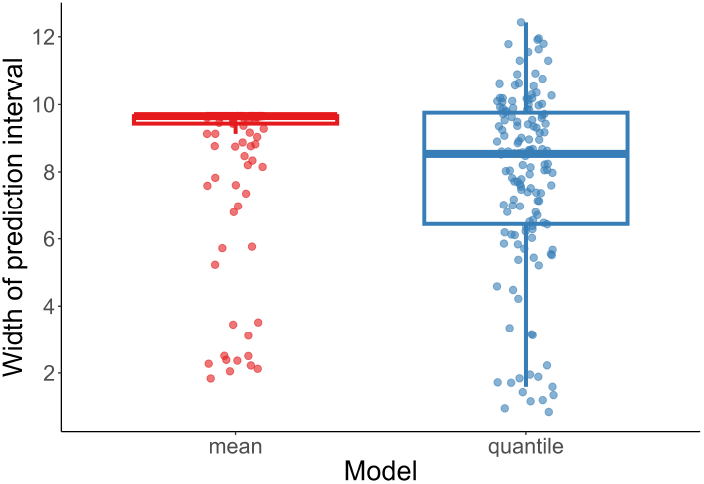
Boxplots of width (unit in year) distribution of prediction intervals derived from conformalized mean regression and conformalized quantile regression methods. The red line refers to the relatively consistent width of prediction intervals derived from conformalized mean regression. The blue boxplot denotes the width distribution of prediction intervals derived from conformalized quantile regression. Clearly, conformalized quantile regression method presents a narrower prediction intervals than conformalized mean regression, demonstrating its improved statistical efficiency.

### 3.5 Prediction intervals with adaptive width better reveals aging pattern

To further investigate the association of the width of prediction intervals with age acceleration, we integrate the average age acceleration (stratified by chronological age) and width of prediction intervals into one plot, as shown in Figure 6. First, we observe that the average acceleration in conformalized mean regression (Panel (a)) is greater than that in conformalized quantile regression (Panel (b)). This implies that the median regression based epigenetic clock fits better into the data, and the corresponding age acceleration better captures the true biological differences rather than measurement error. In addition, both mean and median based methods reveal similar aging patterns: epigenetic age acceleration is the greatest in mid-childhood, and there is a phenomenon of age deceleration in adolescence. However, in contrast to the approximately fixed width intervals constructed via the conformalized mean regression, the width of prediction intervals produced by the quantile regression has a varying pattern that mimics the variation in age acceleration. Such prediction intervals with widths synchronized to biological age acceleration provide important scientific insights: the possible range of epigenetic age varies greatly for children under different developmental stages. Specifically, for children in their mid-childhood and adolescence, the range of their epigenetic ages may be larger than those in early childhood. In particular, the center of the prediction interval in mid-childhood may be slightly higher than the corresponding chronological age while it is the opposite for those in adolescence.

**FIGURE 6.**
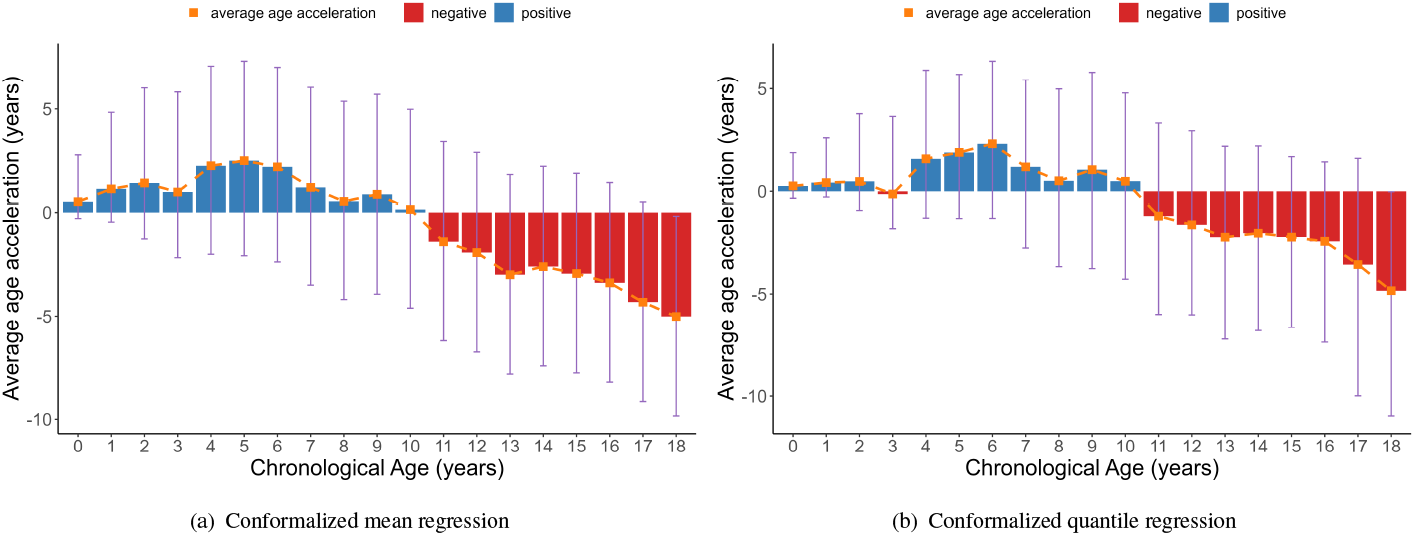
Histogram of the mean value distribution of the age acceleration for individuals in the test sets (age unit in year). The x-axis shows chronological ages, while the y-axis represents the average value in age acceleration. The red column denotes negative value that corresponds to age deceleration, while the blue bar denotes positive average value that corresponds to age acceleration. The orange square indicates the average age acceleration, i.e., predicted values minus the true chronological age, while the purple error bars represent the range of the average age acceleration, i.e., upper and lower bounds of the prediction intervals minus the true chronological age.

## 4 DISCUSSION

Although various epigenetic clocks have been developed to estimate the biological age of an individual based on cellular DNA methylation, their models focus on point prediction only ^1,2^. However, a single point prediction of biological age is not sufficient for clinical decision-making because it does not convey a level of confidence for predicting the age. In this paper, we extend the conformal inference framework to quantify the uncertainty in age prediction, and integrate it with our newly proposed epigenetic clocks developed with high-dimensional quantile regression model with elastic net penalty. We employ data profiled on Illumina 27K and Illumina 450K array platforms to construct a child-specific epigenetic clock that covers the entire period of childhood (0-18 years old). Our prediction and inference framework has the following advantages over existing age prediction models: (a) it comprehensively reflects the heterogeneity in aging patterns across different subpopulations; (b) it provides prediction intervals that will cover the ground truth with certain probability, and thus can enhance the interpretability in practice; (c) the prediction intervals are adaptive to individual DNAm profile and their varying widths uncover the underlying heterogeneity in age acceleration; and (d) the widths derived from the quantile regression models are generally narrower than those from the mean regression models, and thus are statistically more efficient.

Lifespan dynamics in epigenetic biomarkers are rapidly evolving, with significant implications for aging research, personalized medicine, and public health. Birth to late adolescence, for example, is known to be a tremendously dynamic period of development and growth ^40^. It is critical to build biological age prediction models that uncover the underlying population heterogeneity. In our study, we adopt the SIS method to first select the relevant predictors for age prediction in our paper. As an alternative, we also explore variance filtering, which selects features based solely on their variance. However, when we apply the same number of features selected by variance filtering in the mean regression model, its prediction performance is worse than the SIS method, with a correlation of 0.842 and MAE of 24 months. While mean regression has been widely used due to its simplicity and ease of interpretation, it focuses on predictions about the mean outcome. In contrast, quantile regression provides a more detailed picture by estimating the effects of DNAm across the entire distribution of the chronological age, making it especially useful when understanding the impacts on different developmental stages across lifespan is important. Childhood and adolescence are unique periods of rapid change that is unlikely to mimic adulthood in methylome dynamics ^41,42^. Given the differences in the pace of developmental and age-related changes across the life course, developing a unifying prediction model that quantifies the effects of DNAm across different chronological ages will provide a more comprehensive understanding of this dynamic relationship. Our proposed method, when applied to children-specific DNAm datasets, uncovers a clear heterogeneous pattern in different stages of childhood: age acceleration in mid-childhood is much greater than that in toddlerhood, and there is a trend of decceleration in adolescence.

The width of the interval reflects the level of uncertainty associated with the age prediction. Wider intervals indicate higher uncertainty while narrower intervals suggest more confidence in the prediction. This uncertainty can be influenced by various factors such as the inherent noise in the data, the model predictive performance, and the distribution of the data points. According to Figure 2, we observe larger deviations in data points from mid-childhood and adolescence in comparison to those from toddlerhood. This may explain why the resulting intervals in Figure 4 are wider in those two groups to accommodate the increased uncertainty. Such a high variability uncovers the dynamics of DNA methylation levels during mid-childhood, as this period involves significant biological and developmental changes that can be influenced by environmental exposures, genetic factors and lifestyle ^3,43^. While a prediction interval with a width of 10 years seems to be less informative in terms of age prediction, it offers insights on a possible range of DNAm ages in a specific developmental stage. In general, epigenetic age acceleration has been used as a measure of biological aging rate and has been linked to various clinical traits such as physical capability and cognitive functioning ^44^. An age acceleration of 8 years, for example, should be interpreted differently for a person in mid-childhood versus in toddlerhood. Because for the former, such a large deviation is still within its 90% confidence range. From this perspective, our newly proposed pipeline for constructing prediction intervals provides a normal range in which the observed difference between DNAm age and chronological age may arise from dynamics in that specific developmental stage, rather than a systematic indicator of healthy status.

Although our proposed quantile regression based inference framework can be applied to other types of continuous outcomes such as risk of disease or time to death, we currently do not have data on these outcomes to explore whether there will be new insights when the outcome of interest is not chronological age. For example, using quantile regression, we can estimate how DNAm affect different quantiles of the cardiovascular disease risk distribution. The prediction intervals may have varying lengths for those in lower, median and higher risk groups. In conclusion, constructing prediction intervals based on quantile regression provides a nuanced understanding of how DNAm levels and other demographic features influence outcomes of interest across the entire distribution. By leveraging our newly proposed framework, researchers can gain deeper insights into the dynamics of disease risk and develop interventions that are tailored to the specific needs of different risk groups.

## Data Availability

All data produced in the present study are available upon reasonable request to the authors.

